# Associations between Alzheimer’s disease polygenic risk scores and hippocampal subfield volumes in 17,161 UK Biobank participants

**DOI:** 10.1101/2020.10.24.20218925

**Authors:** Heidi Foo, Anbupalam Thalamuthu, Jiyang Jiang, Forrest Koch, Karen A. Mather, Wei Wen, Perminder S. Sachdev

**Affiliations:** Centre for Healthy Brain Aging, CHeBA, School of Psychiatry, University of New South Wales Medicine, Kensington, New South Wales 2052, Sydney, Australia; Neuroscience Research Australia, Randwick, New South Wales 2031, Sydney, Australia; Neuropsychiatric Institute, Euroa Centre, Prince of Wales Hospital, Randwick, New South Wales 2031, Sydney, Australia

**Author notes:** Corresponding author: Heidi Foo, Centre for Healthy Brain Aging, CHeBA, School of Psychiatry, UNSW Medicine, Kensington, 2052, Sydney, Australia.

**Keywords:** Hippocampal subfields, Alzheimer’s disease, Polygenic risk score, Ageing

## Abstract

Hippocampal volume is an important biomarker of Alzheimer’s disease (AD), and genetic risk of AD is associated with hippocampal atrophy. However, the hippocampus is not a uniform structure and has a number of subfields, the associations of which with age, sex, and polygenic risk score for AD (PRS_AD_) have been inadequately investigated. We examined these associations in 17,161 cognitively normal UK Biobank participants (44-80 years). Age was negatively associated with all the hippocampal subfield volumes and females had smaller volumes than men. Higher PRS_AD_ was associated with lower volumes in the bilateral whole hippocampus, hippocampal-amygdala-transition-area (HATA), and hippocampal tail; right subiculum; left cornu ammonis (CA)1, CA4, molecular layer, and granule cell layer of dentate gyrus (CG-DG), with associations being greater on the left side. Older individuals (median age 63 years, n=8984) showed greater subfield vulnerability to high PRS_AD_ compared to the younger group (n=8177), but the effect did not differ by sex. The pattern of subfield involvement in relation to the PRS_AD_ in community dwelling healthy individuals sheds additional light on the pathogenesis of AD.

## 1. Introduction

Hippocampal atrophy is one of the most validated and widely used biomarkers of Alzheimer’s disease (AD) (de Flores et al., 2015). However, the hippocampus is not a homogenous structure and differential vulnerability of the hippocampal subfields to neurodegeneration has been reported to affect different cognitive functions (Mueller et al., 2010). Decline in volumes of hippocampal subfields have been associated with age and sex. Studies in healthy adults have shown significant age effects in CA1-2 (Daugherty et al., 2016; Mueller et al., 2007; Pini et al., 2016; Shing et al., 2011; Wisse et al., 2014), dentate gyrus (DG) (Daugherty et al., 2016; Wisse et al., 2014), and CA4 volumes (Wisse et al., 2014). Evidence has been presented for an anterior-posterior gradient in hippocampal volume reduction (Malykhin et al., 2017) with age, which implies that the hippocampal subregions are not uniformly affected along the hippocampal axis. Significant decreases in cornu ammonis (CA) 1 and subiculum volumes and shape have been observed in participants with mild cognitive impairment (MCI) and AD (de Flores et al., 2015; Pini et al., 2016). Moreover, sex differences in hippocampal subfield volumes have also been observed – males had larger parasubiculum, fimbria, hippocampal fissure, and presubiculum - whereas females had larger volumes for the hippocampal tail (van Eijk et al., 2020). It has also been shown that total hippocampal volume loss in females was more pronounced than males (Nobis et al., 2019). However, to date, there is a lack of studies examining sex differences of hippocampal subfield volumes in greater detail. Therefore, it is important to further study the impact of normal ageing and sex on hippocampal subregions and to determine whether AD genetic risk selectively and differentially impacts distinct subfields.

While one heritability study using twins by Elman et al. (2019) showed little genetic impact on subfields after accounting for total hippocampus volume, a recent genome-wide association study (GWAS) on hippocampal subfield volumes derived from FreeSurfer reported that genetic determinants varied across different subregions (van der Meer et al., 2018). The authors found that the volumes of all hippocampal subfields were heritable in individuals with a mean age of approximately 47.8 (standard deviation = 17.3) years (h^2^ range: 0.14 – 0.27). They identified genome-wide significant single nucleotide polymorphisms (SNPs) associated with the whole hippocampus, presubiculum, subiculum, CA1, dentate gyrus, molecular layer, and hippocampal tail. In addition, in their follow-up analyses on age-stratified subsamples, they also investigated the genetic overlap with AD and showed that three of the significant SNPs were associated with hippocampal subfield volumes in the older age group. This study showed that the differences in cytoarchitecture of the hippocampal subfields are partially driven by genetic variation and that AD-related genes may influence the hippocampal volume predominantly later in life.

Previous studies have identified several common risk variants for AD beyond the well-established apolipoprotein E epsilon 4 allele (*APOE-ε4*) (Li et al., 2018; Xiao et al., 2017). However, these risk alleles have shown small effects on disease risk (Desikan et al., 2015; Lambert et al., 2013). Other studies have used the AD polygenic risk score (PRS_AD_) to examine the cumulative genetic risk for AD (Dezhina et al., 2018; Li et al., 2018), which has shown improvements in predicting cognitive decline beyond the *APOE* locus (Escott-Price et al., 2015).

As hippocampal subfield volumes are differentially affected in the early stages of AD (Zhao et al., 2019), it is possible that this is driven by the genetic risk for AD. Moreover, the PRS_AD_ may have a differential effect on the subfield volumes in a cohort of non-demented individuals. To date, there has not been any study to investigate the relationship between PRS_AD_ and hippocampal subregions, which could help understand the relationship between AD genetic risk factors, ageing, and the pathobiology of AD.

The overall aim of this this study was to investigate hippocampal volumes subfields in greater detail in a large cohort using the UK Biobank data of n = 17,161 participants. The study had three main objectives: (1) to investigate the relationships between age, sex, and hippocampal subfield volumes; (2) to examine the effects of PRS_AD_ constructed using genome-wide significant AD risk variants (Lambert et al., 2013) on hippocampal subfield volumes; (3) to study the interaction of age and sex with the effects of PRS_AD_. We hypothesised that age and sex would be associated with hippocampal subfield volumes. We also hypothesised that there would be varying effects of PRS_AD_ across the different hippocampal subfield volumes, with higher genetic risk of AD being differentially associated with lower hippocampal subfield volumes. We also hypothesised that PRS_AD_ would be modulated by age and sex on hippocampal subfield volumes.

## 2. Materials and methods

### 2.1. Participants

The UK Biobank, which is a large prospective cohort study, included participants from the United Kingdom aged between 40 and 80 years old (Sudlow et al., 2015). Participants provided full informed consent to participate in the UK Biobank and ethics approval was given by the National Health Service National Research Ethics Service (Ref 11/NW/0382).

For this study, cross-sectional data released in January 2019 was used. UK Biobank provided 19,363 participants who had T1-weighted magnetic resonance imaging (MRI) data. Image preprocessing was successful for only 19,275 participants. After quality control filtering and including those with complete brain imaging, genetics, and cognitive data, a final sample of 17,161 participants of European descent was included (see sections 2.2 and 2.3 for more details).

### 2.2. Imaging acquisition and preprocessing

Briefly, the UK Biobank structural T1-weighted MRI scans were acquired on three 3T Siemens Skyra MRI scanners (software platform VD13) at three sites (Reading, Newcastle, and Manchester) using a 32-channel receiving head coil and a 3D MPRAGE protocol (1.0 x 1.0 x 1.0 mm resolution, matrix 208 x 256 x 256, inversion time (TI)/repetition time (TR) = 880/2,000 ms, in-plane acceleration 2). An extensive overview of the data acquisition protocols and image processing carried out on behalf of the UK Biobank can be found elsewhere (Alfaro-Almagro et al., 2018; Miller et al., 2016).

Using the structural T1-weighted MRI images, cortical reconstruction and volumetric segmentations were performed with FreeSurfer v6.0.0 (https://surfer.nmr.mgh.harvard.edu) in our lab. The technical details have been described previously (Fischl & Dale, 2000). Briefly, image processing steps included: motion correction, average of multiple volumetric T1-weighted images, removal of non-brain tissues, automated Talairach transformation, intensity normalisation, segmentation of subcortical white and deep grey matter volumetric structures, tessellation of the grey and white matter boundary, automated topological correction, and surface deformation to optimally place the grey/white and grey/cerebrospinal fluid boundaries (Fischl & Dale, 2000).

Hippocampal subfields were segmented using a Bayesian inference approach and novel atlas using ultra-high resolution (∼0.1 mm isotropic) ex vivo data from autopsy brains (Iglesias et al., 2015). The left and right hippocampi were further segmented into twelve subregions namely, CA1, CA2-3, CA4, fimbria, granule cell layer of dentate gyrus (GC-DG), hippocampus-amygdala-transition-area (HATA), molecular layer, hippocampal fissure, hippocampal tail, parasubiculum, presubiculum, and subiculum. MRI sequences applied in UK Biobank have been designed by imaging experts to optimise the quality of images (Miller et al., 2016). A machine learning-based quality control toolbox was also applied to T1-weighted scans to exclude those with poor acquisition quality (Alfaro-Almagro et al., 2018). To date, there is a lack of standardised quality control method for subcortical regions. Therefore, for quality control, we first examined the principal component (PCA) plots of the sample and omitted any participants who were above 3 standard deviations for the subfields, total hippocampal volumes, and intracranial volume (ICV).

Given that the hippocampal subfields may also not be uniformly affected along the hippocampal axis, we further investigated this using FreeSurfer v7.1.0. (https://surfer.nmr.mgh.harvard.edu/fswiki/HippocampalSubfieldsAndNucleiOfAmygdala). This segmentation automatically partitions the hippocampus into head, body, and tail. This parcellation may also be useful to address the issues related the reliability of smaller hippocampal subfield volumes using FreeSurfer v6.0.0.

### 2.3. Genotyping and Imputation

DNA was extracted from stored blood samples that had been collected from participants on their visit to a UK Biobank assessment centre (Bycroft et al., 2018). The genetic data were acquired using two closely related custom arrays: Affymetrix UK Biobank Lung Exome Variant Evaluation (UK BiLEVE) Axiom array or Affymetrix UK Biobank Axiom array (Bycroft et al., 2018). Quality control was performed using the UK Biobank pipeline. Imputed data set was made available where the UK Biobank interim release was imputed to a reference set combining the UK10K haplotype and 1000 Genome Phase 3 reference panels. For more details, refer to http://biobank.ctsu.ox.ac.uk/crystal/refer.cgi?id=157020. Ten principal components generated by the UK Biobank were included as covariates in the statistical model to control for population stratification.

### 2.4. Polygenic risk score (PRS) derivation

PRS are calculated as a weighted sum of risk alleles an individual carries for any particular disease or phenotype. The weights are the effect sizes observed in GWAS of the relevant phenotype (Choi, Mak, & O’Reilly, 2020). For the current study, PRS derivations were based on the summary statistics from a previous AD GWAS (Lambert et al., 2013). Linkage disequilibrium pruning was performed using the clumping option (r^2^ > 0.2) and a physical distance threshold of 250 KB using the R package, PRSice2 (Euesden et al., 2015). We selected an a-priori threshold of p-value = 5 × 10^−8^, because a previous study has shown that strongest effect for this threshold in predicting risk associated with age at AD onset and that the strength of association was weaker with less stringent PRS thresholds (Leonenko et al., 2019).

The resulting PRS thresholds were z-transformed. As the variants around the *APOE* locus are well-known to be associated with AD, we additionally examined if the effects observed between PRS_AD_ and hippocampal subfield volumes were purely due to the SNPs in *APOE* locus by excluding all the SNPs in this region Chr19:45,116,911-46,318,605 (GRCh37).

### 2.5. Statistical analysis

All statistical analyses were performed using R version 3.5.1 software (The R Foundation, Vienna, Austria). To achieve normality of the skewed hippocampal subfield volumes data, a rank based inverse normal transformation was applied using the R package RNOmni (McCaw, Lane, Saxena, Redline, & Lin, 2019).

Linear regression models were also estimated to study the effects of PRS_AD_, and PRS_AD_ without *APOE* genotype on hippocampal subfield volumes. False Discovery Rate (FDR) adjusted p-values were obtained by using Benjamini & Hochberg (1995) procedure as implemented in the R function *p.adjust*. Significance level was set at adjusted p < 0.05.

In addition, the sample was split into PRS quartiles to investigate differences among the four PRS groups. The fourth quartile (high PRS_AD_, n = 4293) was used as the reference risk group and compared to each of the other three quartiles with the reference group. We further investigated differential influence of PRS_AD_ in relation to age and sex on hippocampal subfield volumes by splitting the cohort into two groups by its median age of 63 years. Those at and below 63 years were referred to as the younger group (n=8984) and those above 63 years as the older group (n=8177).

Age, age^2^, sex, age × sex (i.e. age and sex interaction), age^2^ × sex (i.e. age^2^ and sex interaction), ICV, head position in the scanner, and ten genetic principal components were included in all the analyses as covariates. The imaging and genetic covariates were based on previous literature using the UK Biobank (Elliott et al., 2018).

We also examined the age mediation effect of PRS on the hippocampal subfields. The mediation analysis was done using the R package, mediation (Tingley et al., 2014) and the *p*-values were obtained based on 100000 simulations.

## 3. Results

### 3.1. Age-sex differences in hippocampal subfield volumes

As shown in Table 1, of the 17,161 total participants, 9256 (53.9%) were female and the mean (standard deviation) age was 62.39 (7.43) years. Table 1 shows the hippocampal subfield volumes, showing no significant differences between right and left hemispheres.

**Table 1.** UK biobank sample characteristics and mean (standard deviation) of hippocampal subfield volumes in mm^3^

| UK Biobank sample characteristics |  |  |  |  |
| --- | --- | --- | --- | --- |
| Sample size | 17,161 |  |  |  |
| Age (mean, SD) | 62.39 (7.43) years |  |  |  |
| Gender (n, %) | 9256 (53.9%) females; 7905 (46.1%) males |  |  |  |
| Hippocampal subfields | Left hemisphere | Right hemisphere | *t* | *p* |
| CA1 | 637.09 (75.61) | 666.69 (79.81) | -0.095 | 0.924 |
| CA2-3 | 214.18 (30.23) | 237.29 (32.43) | -0.239 | 0.812 |
| CA4 | 260.59 (29.34) | 276.30 (31.72) | 0.164 | 0.870 |
| Subiculum | 436.56 (50.21) | 438.32 (49.54) | 0.140 | 0.893 |
| Presubiculum | 307.85 (38.70) | 297.23 (36.42) | -0.061 | 0.951 |
| Parasubiculum | 64.97 (13.14) | 62.89 (12.17) | -0.072 | 0.943 |
| Hippocampal tail | 519.89 (70.64) | 560.88 (72.71) | -0.025 | 0.980 |
| Hippocampal fissure | 170.80 (31.39) | 176.18 (32.44) | 0.277 | 0.781 |
| HATA | 61.29 (9.78) | 66.00 (9.97) | 0.212 | 0.832 |
| Molecular layer | 573.55 (62.41) | 592.21 (64.70) | 0.012 | 0.990 |
| GC-DG | 302.41 (34.23) | 318.72 (36.45) | 0.093 | 0.926 |
| Fimbria | 79.06 (18.60) | 73.11 (17.11) | -0.659 | 0.510 |
| Whole hippocampus | 3457.44 (351.80) | 3589.66 (365.72) | -0.041 | 0.967 |

Abbreviations: CA, cornu ammonis; HATA, hippocampus-amygdala-transition-area; GC-DG, granule cell layer of dentate gyrus. Significant differences were assessed between left and right hippocampal sub-fields using t-tests.

All hippocampal subfield volumes were negatively associated with age (Supplementary table 1, Supplementary figure 1). Sex was associated with all hippocampal subfield volumes, except for the left hippocampal tail, with women having lower hippocampal volumes than men in all regions (Supplementary table 1, Supplementary figure 1). Age and sex interactions were also negatively associated with all the hippocampal subfield volumes, except left CA2-3, left CA4, left parasubiculum, and right presubiculum (Supplementary table 1). Taken together, age and sex were significantly associated with the hippocampal subfield volumes.

### 3.2. Associations between PRS_AD_ and hippocampal subfield volumes

Table 2 summarises the associations between PRS_AD_ and hippocampal subfield volumes. Left and right hippocampal volumes were negatively associated with PRS_AD_. In addition, hippocampal subregions, namely bilateral HATA, bilateral hippocampal tail, right subiculum, left CA1, left CA4, left molecular layer, and left CG-DG, were also found to be inversely associated with PRS_AD_. The results imply that higher genetic load for AD was related to lower hippocampal subfield volumes. Summary statistics of the regression model corresponding to all the subfield volumes are presented in the supplementary table 1. When we repeated these analyses controlling for total hippocampal volume in addition to other covariates, the following subfields had significant associations (unadjusted p-value < 0.05) namely, right hippocampal fissure, left HATA, left presubiculum, and left fimbria. However, after FDR adjustment, none of these associations was statistically significant (Supplementary Table 2f).

**Table 2.** Associations between hippocampal subfield volumes and PRS_AD_ (threshold 5 x10^−8^)

| Hippocampal subfields | $\beta$ | SE | $t$ | $p$ | *Adjusted p-value* |
| --- | --- | --- | --- | --- | --- |
| Left CA1 | -0.0209 | 0.0061 | -3.4196 | 0.000629 | 0.005448* |
| Right CA1 | -0.0112 | 0.0062 | -1.8230 | 0.068324 | 0.118428 |
| Left CA2-3 | -0.0130 | 0.0067 | -1.9260 | 0.054116 | 0.100501 |
| Right CA2-3 | -0.0087 | 0.0065 | -1.3361 | 0.181525 | 0.262203 |
| Left CA4 | -0.0160 | 0.0063 | -2.5402 | 0.011088 | 0.028828* |
| Right CA4 | -0.0098 | 0.0062 | -1.5664 | 0.117263 | 0.179343 |
| Left fimbria | 0.0063 | 0.0069 | 0.9130 | 0.361248 | 0.494339 |
| Right fimbria | 0.0015 | 0.0069 | 0.2107 | 0.833092 | 0.902516 |
| Left GC-DG | -0.0180 | 0.0061 | -2.9459 | 0.003225 | 0.011978* |
| Right GC-DG | -0.0108 | 0.0061 | -1.7604 | 0.078356 | 0.127328 |
| Left HATA | -0.0254 | 0.0068 | -3.7449 | 0.000181 | 0.003688* |
| Right HATA | -0.0213 | 0.0067 | -3.1799 | 0.001476 | 0.009595* |
| Left hippocampal fissure | -0.0046 | 0.0069 | -0.6652 | 0.505960 | 0.657749 |
| Right hippocampal fissure | 0.0003 | 0.0067 | 0.0496 | 0.960448 | 0.960448 |
| Left hippocampal tail | -0.0203 | 0.0070 | -2.8890 | 0.003869 | 0.012574* |
| Right hippocampal tail | -0.0250 | 0.0069 | -3.6305 | 0.000284 | 0.003688* |
| Left molecular layer | -0.0179 | 0.0059 | -3.0264 | 0.002479 | 0.010742* |
| Right molecular layer | -0.0120 | 0.0059 | -2.0313 | 0.042240 | 0.084480 |
| Left parasubiculum | -0.0027 | 0.0070 | -0.3859 | 0.699589 | 0.826787 |
| Right parasubiculum | -0.0036 | 0.0063 | -0.5740 | 0.565988 | 0.700747 |
| Left presubiculum | -0.0017 | 0.0064 | -0.2625 | 0.792912 | 0.896335 |
| Right presubiculum | 0.0004 | 0.0070 | 0.0592 | 0.952767 | 0.960448 |
| Left subiculum | -0.0128 | 0.0062 | -2.0469 | 0.040680 | 0.084481 |
| Right subiculum | -0.0156 | 0.0063 | -2.4834 | 0.013024 | 0.030785* |
| Left whole hippocampus | -0.0180 | 0.0058 | -3.1134 | 0.001853 | 0.009633* |
| Right whole hippocampus | -0.0150 | 0.0058 | -2.5944 | 0.009482 | 0.027393* |

Abbreviations: HATA, hippocampus-amygdala-transition-area; CA1, cornu ammonis; GC-DG, granule cell layer of dentate gyrus. \*Denotes statistically significant results after FDR correction.
Note: This analysis was run with the full regression model (adjusted for age, age2, sex, age × sex (i.e. age and sex interaction), age2 × sex (i.e. age2 and sex interaction), intracranial volume (ICV), head position in the scanner, and ten genetic principal components). The full results can be found in Supplementary table 1.

In addition, when we investigated the effect of PRS_AD_ on hippocampal subfield volumes segmented into head, body, and tail using FreeSurfer v7.1.0, significant results were observed in all the segmented regions including bilateral whole hippocampus (Supplementary Table 3). Given that this is not informative on which regions were primarily affected, using the segmentation from FreeSurfer v6.0.0 may shed more light onto the differential vulnerability of the hippocampus. Therefore, we only report findings using the FreeSurfer v6.0.0 segmentation from here on.

To determine whether the effect of PRS_AD_ on hippocampal subfield volumes was independent of the *APOE* genotype, we looked at the effect of PRS_AD_ without SNPs in *APOE* locus (Supplementary table 4). As most of the SNPs (73%) used for PRS_AD_ were from chromosome 19, the PRS_AD_ without the *APOE* locus was not associated with the hippocampal subfields.

### 3.3. Modulation of PRS_AD_ on hippocampal subfield volumes by age and sex

Higher genetic risk for AD, as specified by comparing the fourth quartile versus the first quartile of the PRS_AD_, was associated with increased risk for lower volume in the left whole hippocampal, bilateral HATA, left CA1, left molecular layer, left CG-DG, and right hippocampal tail compared to those with low PRS_AD_ (OR 1.04 – 1.06, adjusted *p* < 0.05; Supplementary table 5).

In addition, modulation of PRS_AD_ by age in the hippocampal subfield volumes was observed. Risk for volume loss between low and high PRS_AD_ was similar among younger participants (≤ 63 years old) (Supplementary table 6). Older participants were susceptible to lower bilateral HATA, left CA1, and right hippocampal tail volumes in high versus low PRS_AD_ (OR 2.07 – 2.55, adjusted *p* < 0.012) (Supplementary table 7, figure 1).

**Figure 1.**
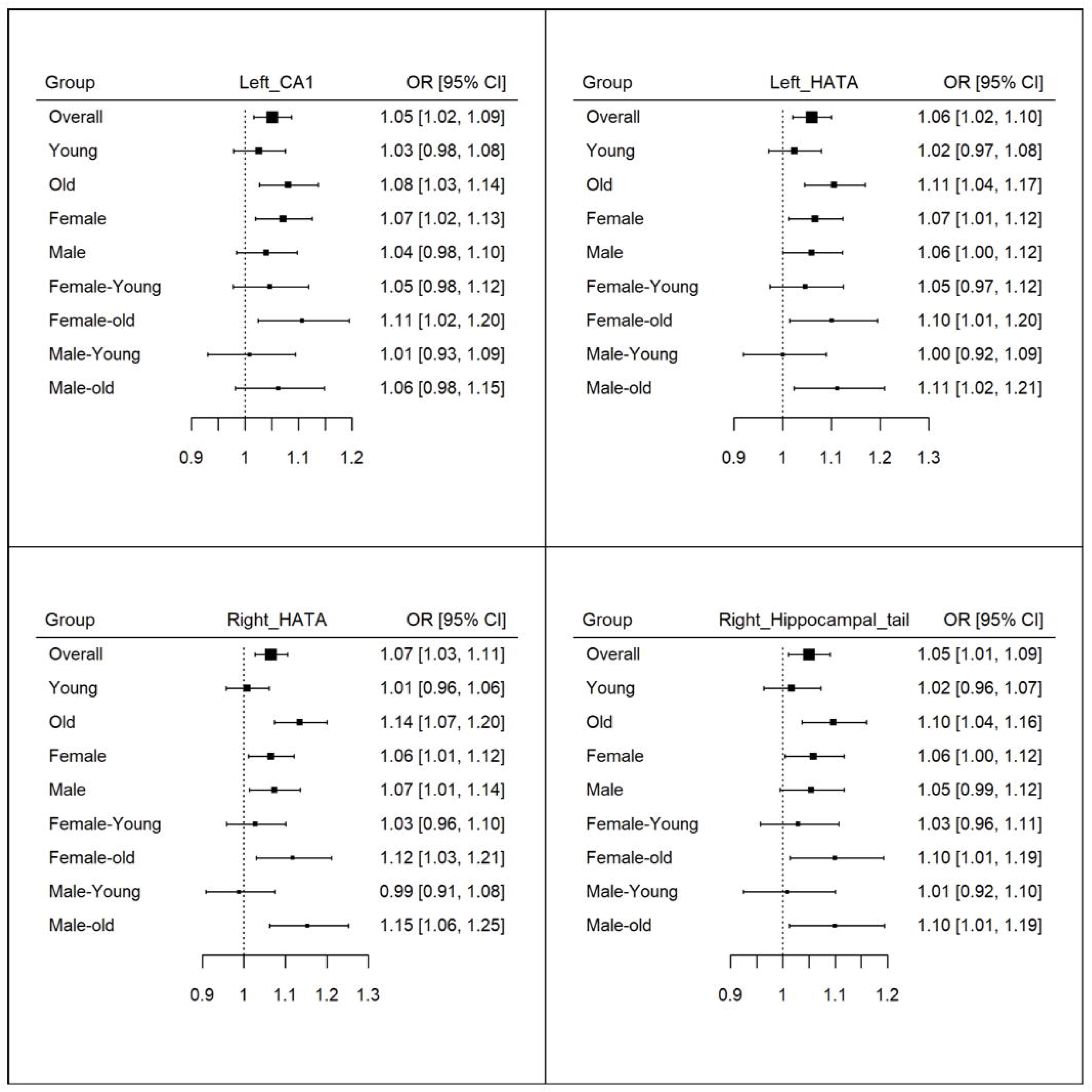
Forest plot with bilateral HATA, left CA1, and right hippocampal tail for all the group comparisons between Q1, first quartile, and Q4, last quartile. Note: The subgroups include overall, young, old, female, male, female-young, female-old, male-young, and male-old.

There were no significant sex differences in the effect of PRS on subfield volumes (Supplementary tables 8-9, Supplementary figure 2). There were also no significant differences in the hippocampal subfield volumes associations with PRS_AD_ when we split the group into quartiles for age and sex (Supplementary tables 10-13).

### 3.4. Mediation of age on the relationship between PRS_AD_ and hippocampal subfield volumes

Given that the findings show age differences in the effect of PRS_AD_ on hippocampal subfield volumes, we further examined whether age mediated this relationship. The mediation analysis showed that age mediated the relationship between PRS_AD_ and hippocampal subfield volumes (Supplementary table 14).

## 4. Discussion

The main findings of this study are: (i) hippocampal subfield volumes are associated with age and sex; (ii) Higher PRS_AD_ is associated with lower hippocampal subfield volumes in the left and right whole hippocampus, bilateral HATA, bilateral hippocampal tail, right subiculum, left CA1, left CA4, left molecular layer, and left CG-DG, which suggests differential effect of PRS_AD_ on the hippocampal subfield volumes; (iii) Modulation of PRS_AD_ by age, but not sex, on hippocampal subfield volumes, with associations seen in older but not younger individuals; and (iv) the associations between PRS_AD_ and hippocampal subfield volumes were driven by the SNPs in the *APOE* locus.

Hippocampal subfield volumes were associated with age. In line with previous studies, we reported that all the volumes of hippocampal subfields decrease with age in healthy adults (Daugherty et al., 2016; Malykhin et al., 2017; Zheng et al., 2018). However, the age-related changes in hippocampal subfields varied across studies – whereas one showed decline of CA1-2 and CA3-DG (Malykhin et al., 2017), another found no difference in these subfields across the human lifespan (Daugherty et al., 2016). In addition, hippocampal subfield volumes were also associated with sex in our study. One study using the UK Biobank data reported that total hippocampal volume loss was more pronounced in females than males (Nobis et al., 2019), which corroborates our findings, although this study was based on overlapping samples. However, few studies have examined sex differences in hippocampal subfields, and the focus has been on CA1, CA3, or dentate gyrus (Duarte-Guterman et al., 2015; Koss & Frick, 2017; McEwen, 2010; Scharfman & MacLusky, 2017). To date, no study has investigated sex differences in hippocampal subfield volumes. Our study is therefore novel in this regard, and we found that females had significantly lower hippocampal volumes than males in most hippocampal subfield volumes, except for the left fimbria, bilateral GC-DG, bilateral CA4, right HATA, and left hippocampal tail, even after controlling for ICV. However, it remains to be elucidated why this might be. More studies are warranted in order to understand this sex effect on hippocampal subfield volumes. Taken together, these results imply that the hippocampus may have differential vulnerabilities to age and sex. Thus, it may be more valuable to study hippocampus subregions rather than as a whole.

Consistent with previous ante- and post-mortem studies, which reported that participants at higher genetic risk of AD had lower CA1 hippocampal volumes compared to age-matched controls (Padurariu et al., 2012; Zhao et al., 2019), we observed that PRS_AD_ was negatively associated with left CA1. Moreover, previous studies have also found that hippocampal subfields, particularly the CA1 and/or subiculum (de Flores et al., 2015), have been reported to be a potential indicator of conversion from MCI to AD, the higher association observed between PRS_AD_ and CA1 may identify individuals at risk of AD. In addition, the dentate gyrus, which shows reduced neurogenesis in AD and ageing (Adler et al., 2018; Hollands et al., 2016), was also observed to be associated with PRS_AD_. This implies that AD genetic risk may be able to predict left CA1 and left dentate gyrus atrophy in healthy adults without overt disease. Confirmed by previous studies reporting that subiculum is involved in early phases of AD, we also showed that subiculum is associated with PRS_AD_ (Carlesimo et al., 2015; Zhao et al., 2019). However, presubiculum and fimbria, previously reported to be associated with AD (Carlesimo et al., 2015; Zhao et al., 2019), were not observed in this study. It is possible that atrophy in these regions occurs later in the course of the disease and may not be sensitive in identifying participants at risk of developing AD. Our findings also showed that PRS_AD_ was associated with bilateral hippocampal tail, bilateral HATA, left CA4, and left molecular layer. While these regions have not been shown to be involved in the trajectory of AD in previous studies, they show genetic effects in the pathophysiology of AD. This finding needs to be replicated in order to understand why these additional subfields might be affected.

We also observed that the effects of PRS_AD_ p-value threshold of 5 × 10^−8^ were mainly driven by the *APOE* genotype. The *APOE* genotype is one of the most commonly studied variants in ageing due to the high odds (14.9:1) for developing AD for ε4 homozygotes (Farrer et al., 1997). Notably, *APOE-ε4* has been associated with greater thinning of hippocampal CA1 (Kerchner et al., 2014). Another longitudinal study demonstrated that greater atrophy of the left than right hippocampus was most pronounced in *APOE-ε4* homozygotes compared to heterozygotes over time (Li et al., 2016). As the attributable fraction to AD risk added by all other polymorphisms besides *APOE* was the only about 1.0%-8.0% while *APOE* accounted for 27.3% (Lambert et al., 2013), it is therefore likely that the effect of other polymorphisms is small and is not seen when examining multiple hippocampal subfield volumes. Consequently, it seems that the *APOE* genotype makes the most salient contribution to this effect.

Our findings also underline that bilateral HATA, left CA1, and right hippocampal tail showed slightly greater vulnerability in the older age group with those with higher PRS_AD_ than lower PRS_AD_. While ageing may negatively affect neuronal homeostasis (Boisvert et al., 2018), it has been observed that the presence of *APOE-ε4* alters normal function of glial cells and may contribute to accelerated ageing and neurodegenerative disease pathogenesis (Fernandez et al., 2019). One study reported that a PRS_AD_ calculated predictability for late-onset AD of 82.5% (where the *APOE* SNPs, rs7412 and rs429358, which determine the ApoE isoforms, alone had predictability of 81.8%) and 61.0% of late-onset AD conversion from mild cognitive impairment (Chaudhury et al., 2019). They further postulated healthy controls with higher PRS may represent those who eventually develop disease if they had lived longer, as the average age at death for controls with higher PRS was significantly reduced than those with lower PRS (Chaudhury et al., 2019). Given that we observed that bilateral HATA, left CA1, and right hippocampal tail were associated with age, sex, and PRS_AD_ in our study, it is possible that changes in these regions may be hallmarks of both normal ageing and AD.

Furthermore, we examined the overlap of the SNPs between the hippocampal subfield GWAS (van der Meer et al., 2018) and in our PRS_AD_ analysis. None of the 60 SNPs used in PRS_AD_ were found among the 23 unique GWAS hits across all subfield volumes. However, when we looked at the full list of 25 GWAS summary statistics for the subfields, we found a few SNPs, including rs80307900 and rs12459810 on chromosome 19, and rs7982 on chromosome 8 associated with multiple hippocampal subfield volumes at suggestive significance (p < 0.05) (Supplementary tables 15-16). This implies that AD genes may differentially affect the hippocampal subfield volumes.

The challenge to understand why several hippocampal subfield volumes were associated with PRS_AD_ remains. Our investigation of PRS_AD_ in a non-demented ageing cohort from the UK Biobank found hippocampal subfields that are affected by genetic risk for AD (subtle AD pathology). The participants in this study are community dwelling individuals but as literature has shown, AD pathology begins 10-20 years before the first clinical manifestations (Hof et al., 1996; Perl, 2010). Such findings are supported by a previous study by Foley et al. (2017), which showed an effect of PRS_AD_ on hippocampal volume that was detected in young adults. These results suggest that AD genetic risk may be associated with hippocampal subfield volume changes that have been implicated in the early stages of AD pathology decades prior to disease onset. It should be noted that further multiple insults, such as cardiovascular disease and environmental factors, may be required for AD pathology to fully develop. Longitudinal follow-up of the UK Biobank participants will reveal those participants that go on to develop dementia and will allow a more detailed examination of this question.

The strengths of this study include the well characterised large sample size cohort and the uniformed MRI and genetic methods. Limitations should also be considered. The cross-sectional nature of the study prevents the ability to detect subtle changes in the hippocampal subfield volumes over time within individuals and those who go on to develop AD cannot be identified. In addition, the inherent nature of polygenic analysis precludes identifying specific biological mechanisms that contribute to the hippocampal subfield differences (Foley et al., 2017). Further, given that there has not been any consensus in regards to the optimal p-value threshold to use for the PRS analysis and the increase of multiple test burden investigating multiple thresholds, future studies might benefit from investigating the optimal p-value threshold to use. Moreover, comparison of results with existing literature is impeded by the variation in the extraction methods used for calculating subfield volumes. For example, some investigators have used FreeSurfer parcellation like in this study, while others use manual segmentation (van der Meer et al., 2018). In addition, due to the reported lower reliability of using volumes from smaller subfields, such as the fimbria, hippocampal fissure, HATA, and parasubiculum (Quattrini et al., 2020; Worker et al., 2018), the findings should be carefully interpreted. Furthermore, although it has been observed that years of education is associated with hippocampal subfield volumes (Kang et al., 2018), the lack of information on the years of education in the UK Biobank precludes us from including it as a covariate.

## 5. Conclusion

This study shows that the genetics of AD, specifically the *APOE* locus, are a contributing factor for the differential hippocampal subfield vulnerabilities seen in non-demented older adults, and the pattern of volume loss seems to be similar to that observed in the early stages of AD. The effect of PRS_AD_ and specific hippocampal subfield volumes may be useful in allowing us to understand the genetic effects on individual subfields. It furthers our knowledge of the association of AD risk with subfields of the hippocampus, with a focus specific to the subfields that have not received detailed investigation. Such fine-grained analysis of the hippocampus is arguably important in trying to understand which regions are most vulnerable to AD pathological mechanisms, and potentially add refinement to the notion of the hippocampus as a biomarker of AD.

## Data Availability

The data is available on the UK Biobank.

https://www.ukbiobank.ac.uk/

## Acknowledgements

Many thanks to the UNSW Scientia PhD Scholarship Program for their support to Foo, H. This research has been conducted using the UK Biobank Resource (Project ID: 37103).

---

Supplementary table 1. Results from base regression model examining PRS_AD_ and hippocampal subfield volumes while controlling for age, age^2^, sex, age × sex, age^2^ × sex, intracranial volume (ICV), head position in the scanner, and ten genetic principal components.

Abbreviations: SE, standard error; CA, cornu ammonis; DG, granule cell layer of dentate gyrus; HATA, hippocampal-amygdala-transition-area

Supplementary table 2. Results from base regression model examining PRS_AD_ and hippocampal subfield volumes while controlling for covariates including total hippocampal volume

Abbreviations: SE, standard error; CA, cornu ammonis; DG, granule cell layer of dentate gyrus; HATA, hippocampal-amygdala-transition-area

Supplementary table 3. Results from base regression model examining PRSAD and hippocampal head, body, and tail volumes segmented using FreeSurfer 7.1.0

Abbreviations: SE, standard error; CA, cornu ammonis; DG, granule cell layer of dentate gyrus; HATA, hippocampal-amygdala-transition-area

Supplementary table 4. Regression model of PRS_AD_ without *ApoE* locus and hippocampal subfield volumes while controlling for age, age^2^, sex, age × sex, age^2^ × sex, intracranial volume (ICV), head position in the scanner, and ten genetic principal components.

Abbreviations: SE, standard error; CA, cornu ammonis; DG, granule cell layer of dentate gyrus; HATA, hippocampal-amygdala-transition-area

Supplementary table 5. Influence of low and high PRS_AD_ on hippocampal subfield volumes Abbreviations: OR, odds ratio; Q1, first quartile; Q2, second quartile; Q3, third quartile; SE, standard error; CA, cornu ammonis; DG, granule cell layer of dentate gyrus; HATA, hippocampal-amygdala-transition-area

Supplementary table 6. Risk for hippocampal subfield volume loss between low and high PRS_AD_ in younger participants (≤ 63 years old)

Abbreviations: OR, odds ratio; Q1, first quartile; Q2, second quartile; Q3, third quartile; Q4all, comparison between fourth quartile and the rest of the quartiles; SE, standard error; CA, cornu ammonis; GC-DG, granule cell layer of dentate gyrus; HATA, hippocampal-amygdala-transition-area

Note: Refer to ORQ1 and pval_prsQ1_adj for hippocampal volumes between low and high PRS_AD_.

Supplementary Table 7. Risk for hippocampal subfield volume loss between low and high PRS_AD_ in older participants (> 63 years old)

Abbreviations: OR, odds ratio; Q1, first quartile; Q2, second quartile; Q3, third quartile; Q4all, comparison between fourth quartile and the rest of the quartiles; SE, standard error; CA, cornu ammonis; GC-DG, granule cell layer of dentate gyrus; HATA, hippocampal-amygdala-transition-area

Note: Refer to ORQ1 and pval_prsQ1_adj for hippocampal volumes between low and high PRS_AD_.

Supplementary table 8. Hippocampal subfield volumes associations with PRS_AD_ split into quartiles for females

Abbreviations: OR, odds ratio; Q1, first quartile; Q2, second quartile; Q3, third quartile; Q4all, comparison between fourth quartile and the rest of the quartiles; SE, standard error; CA, cornu ammonis; GC-DG, granule cell layer of dentate gyrus; HATA, hippocampal-amygdala-transition-area

Note: Refer to ORQ1 and pval_prsQ1_adj for hippocampal volumes between low and high PRS_AD_.

Supplementary table 9. Hippocampal subfield volumes associations with PRS_AD_ split into quartiles for males

Abbreviations: OR, odds ratio; Q1, first quartile; Q2, second quartile; Q3, third quartile; Q4all, comparison between fourth quartile and the rest of the quartiles; SE, standard error; CA, cornu ammonis; GC-DG, granule cell layer of dentate gyrus; HATA, hippocampal-amygdala-transition-area

Note: Refer to ORQ1 and pval_prsQ1_adj for hippocampal volumes between low and high PRS_AD_.

Supplementary tables 10-13. Hippocampal subfield volumes associations with PRS_AD_ split into quartiles for age and sex i.e. females ≤ 63 years (Supplementary table 10), females > 63 years (Supplementary table 11), males ≤ 63 years (Supplementary table 12), males > 63 years (Supplementary table 13).

Abbreviations: OR, odds ratio; Q1, first quartile; Q2, second quartile; Q3, third quartile; Q4all, comparison between fourth quartile and the rest of the quartiles. SE, standard error; CA, cornu ammonis; GC-DG, granule cell layer of dentate gyrus; HATA, hippocampal-amygdala-transition-area

Note: Refer to ORQ1 and pval_prsQ1_adj for hippocampal volumes between low and high PRS_AD_.

Supplementary table 14. Mediation analysis on the effects of age on the relationship between PRS_AD_ and hippocampal subfield volumes.

Abbreviations: direct, direct effect of PRS_AD_ in in the model with the mediator variable; total, effect of PRS_AD_ in the model without the mediator variable; med, mediation effect (total effect - direct effect); CA, cornu ammonis; DG, granule cell layer of dentate gyrus; HATA, hippocampal-amygdala-transition-area

Supplementary tables 15-16. Intersect between hippocampal subfield volumes with bilateral whole hippocampus as covariate (Supplementary Table 15) and bilateral whole hippocampus as independent variable together with other hippocampal subfield volumes (Supplementary Table 16) and PRS_AD_.

Note: Highlighted fields are SNPs that reached suggestive significance of p < 0.05.

Supplementary figure 1. Age and sex trajectory for hippocampal subfield volumes. Quadratic curve was fitted using the residuals adjusted for imaging and principal components. Line represents the fitted curve for age and dots represents the confidence intervals.

Abbreviations: CA, cornu ammonis; DG, granule cell layer of dentate gyrus; HATA, hippocampal-amygdala-transition-area

Supplementary figure 2. Forest plot with hippocampal subfields ROI that were not significant for all the group comparisons between Q1, first quartile, and Q4, last quartile.

Note: The subgroups include overall, young, old, female, male, female-young, female-old, male-young, and male-old.

