## Supplementary figures and images for "Associations between Alzheimer’s disease polygenic risk scores and hippocampal subfield volumes in 17,161 UK Biobank participants"

### Supplementary figure 1

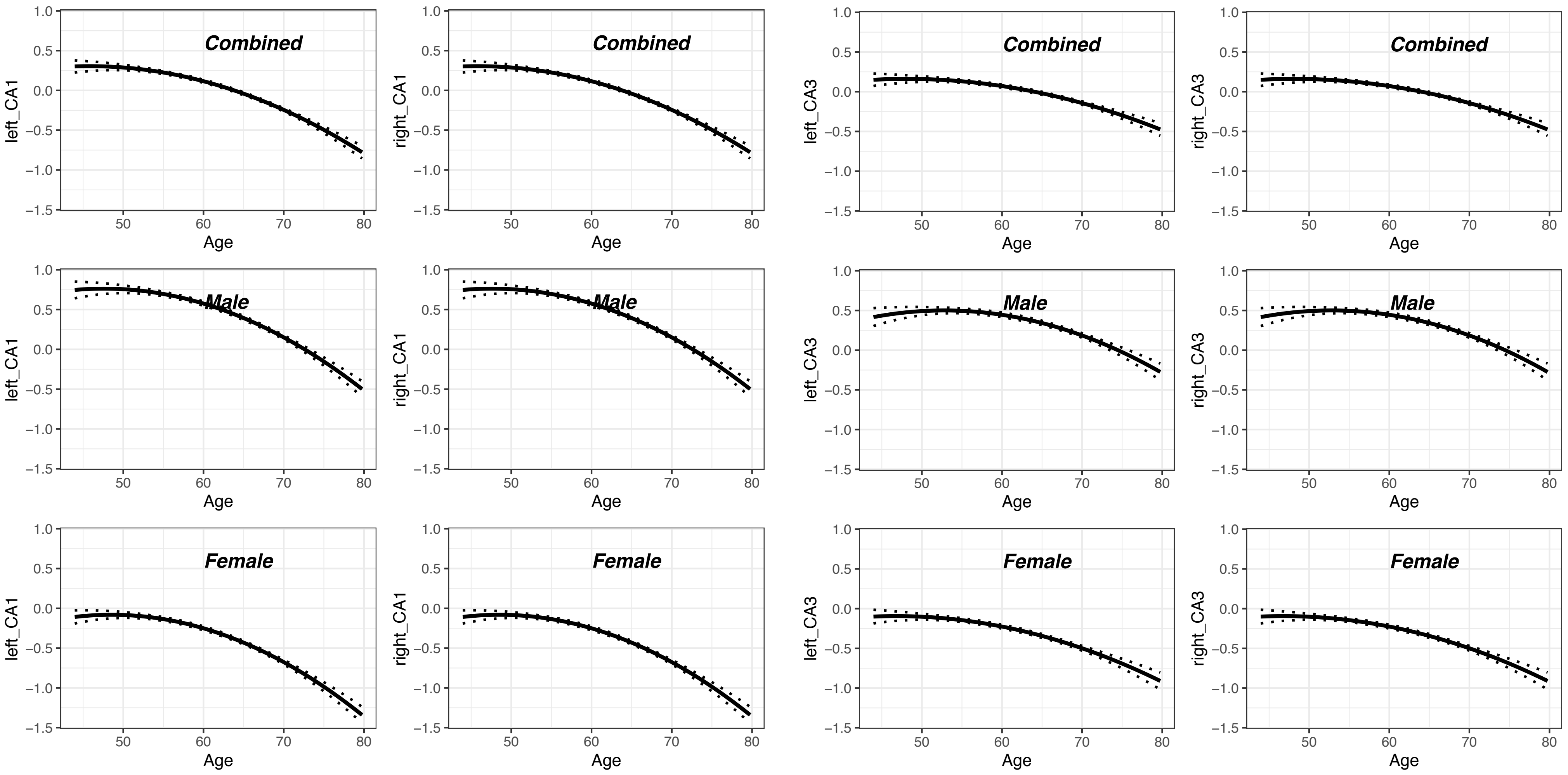

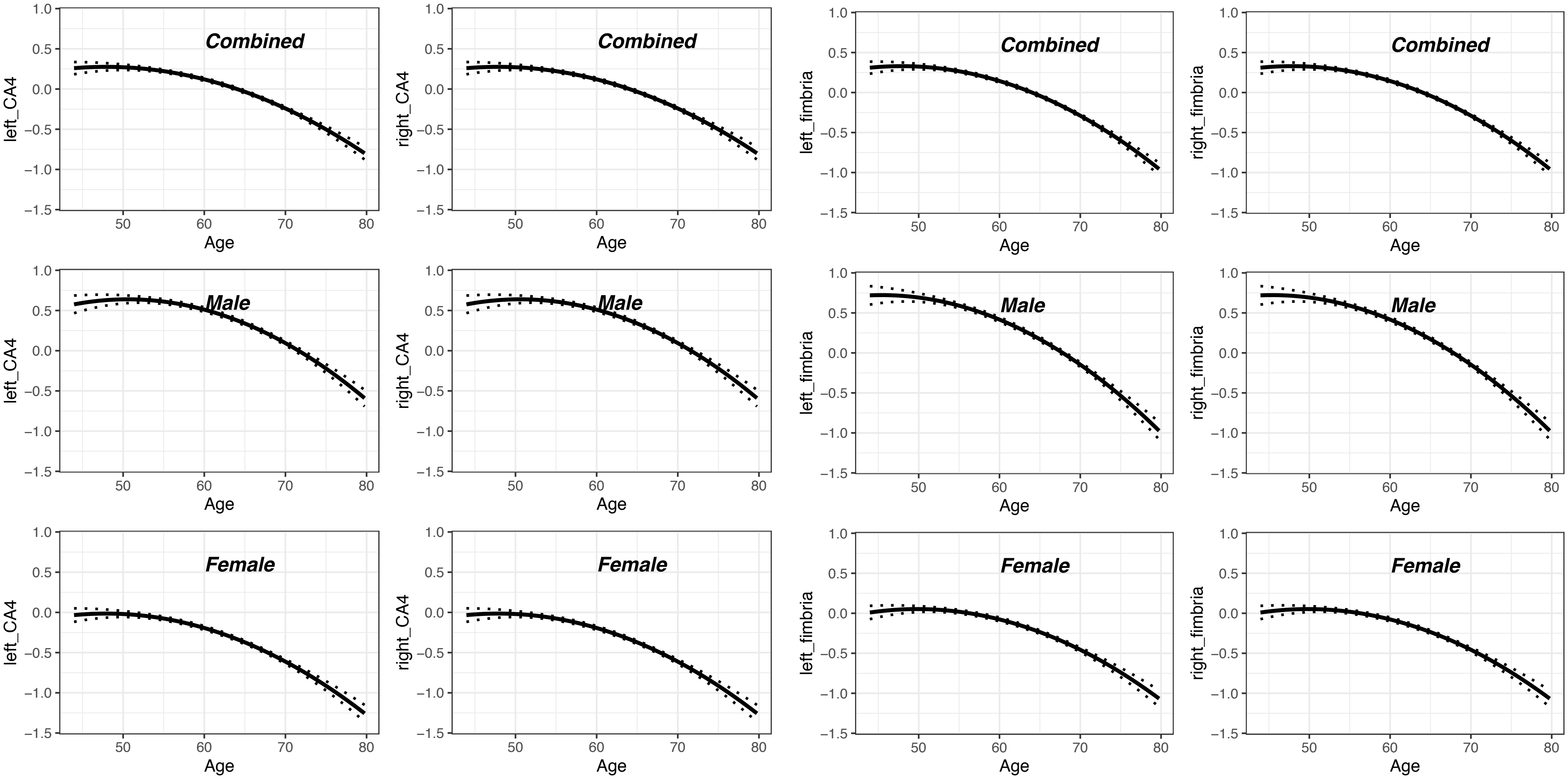

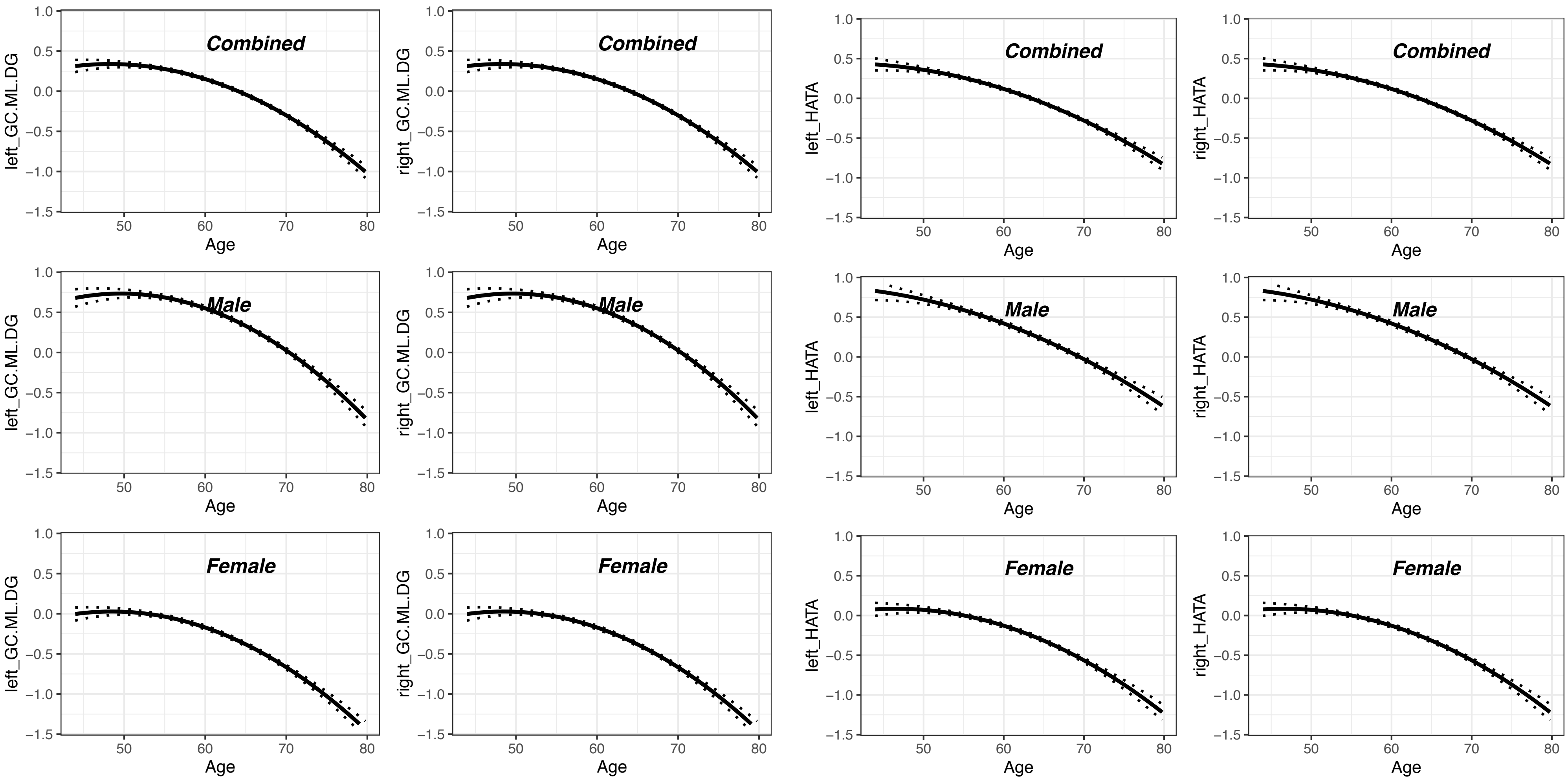

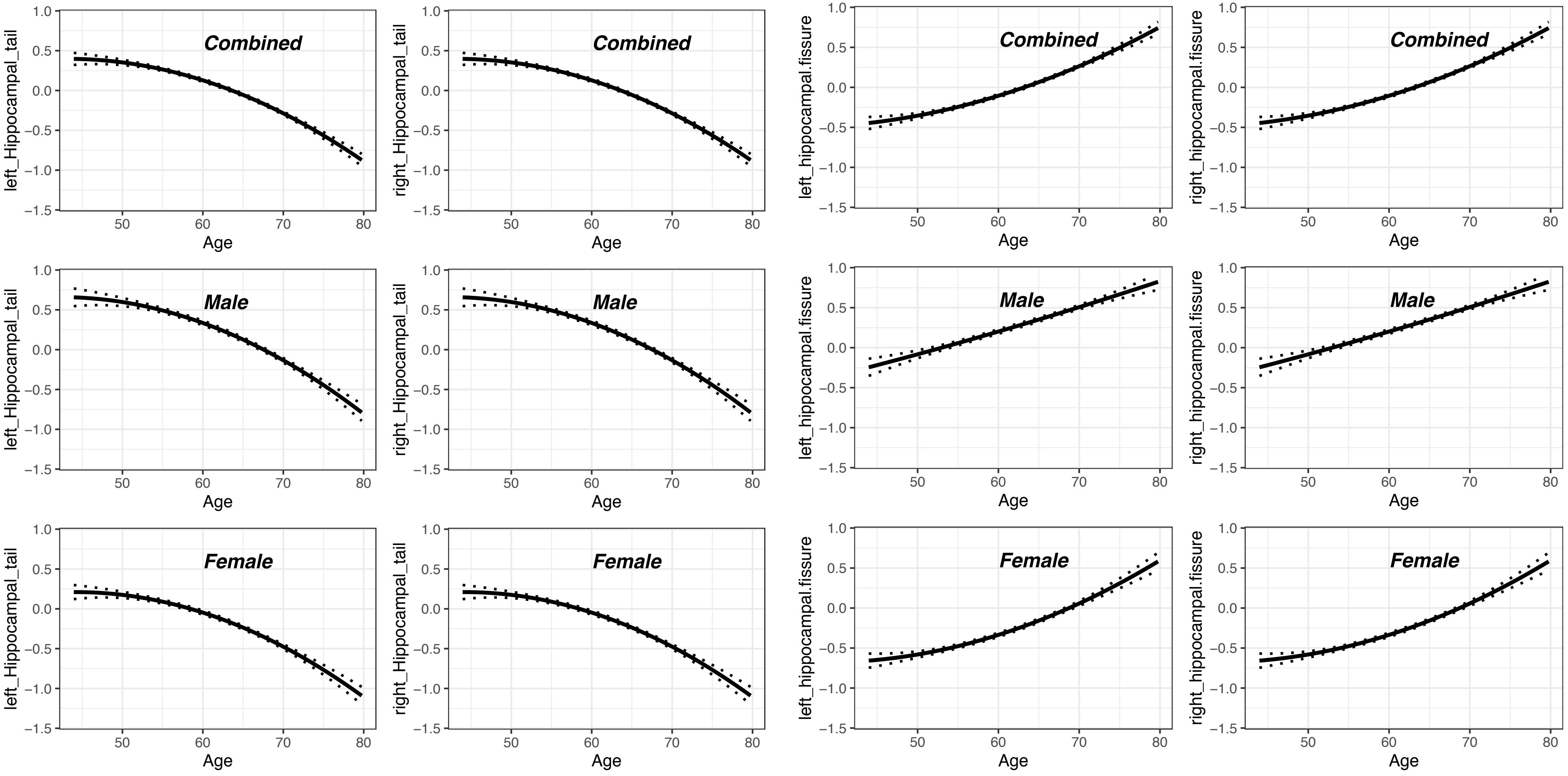

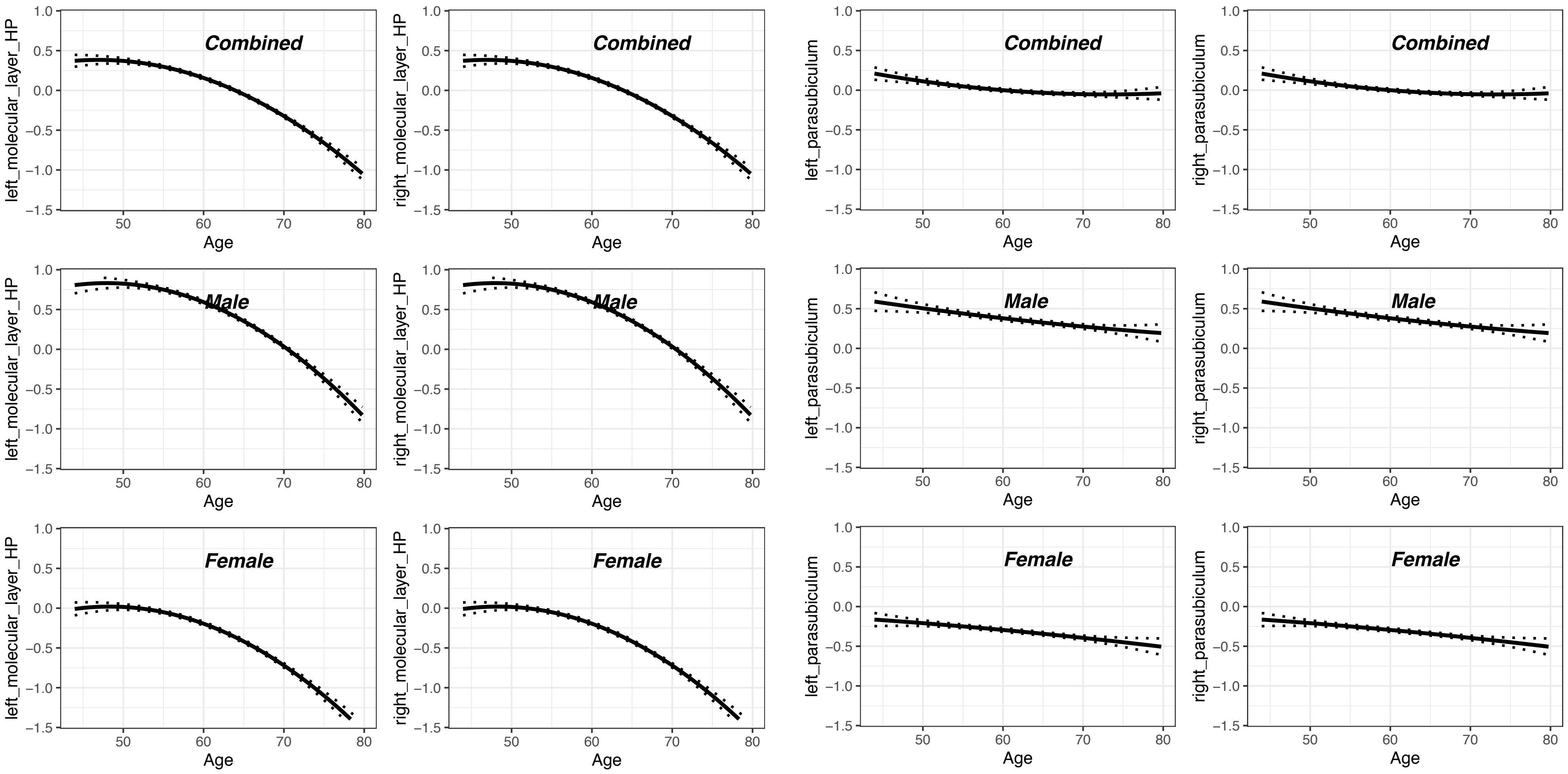

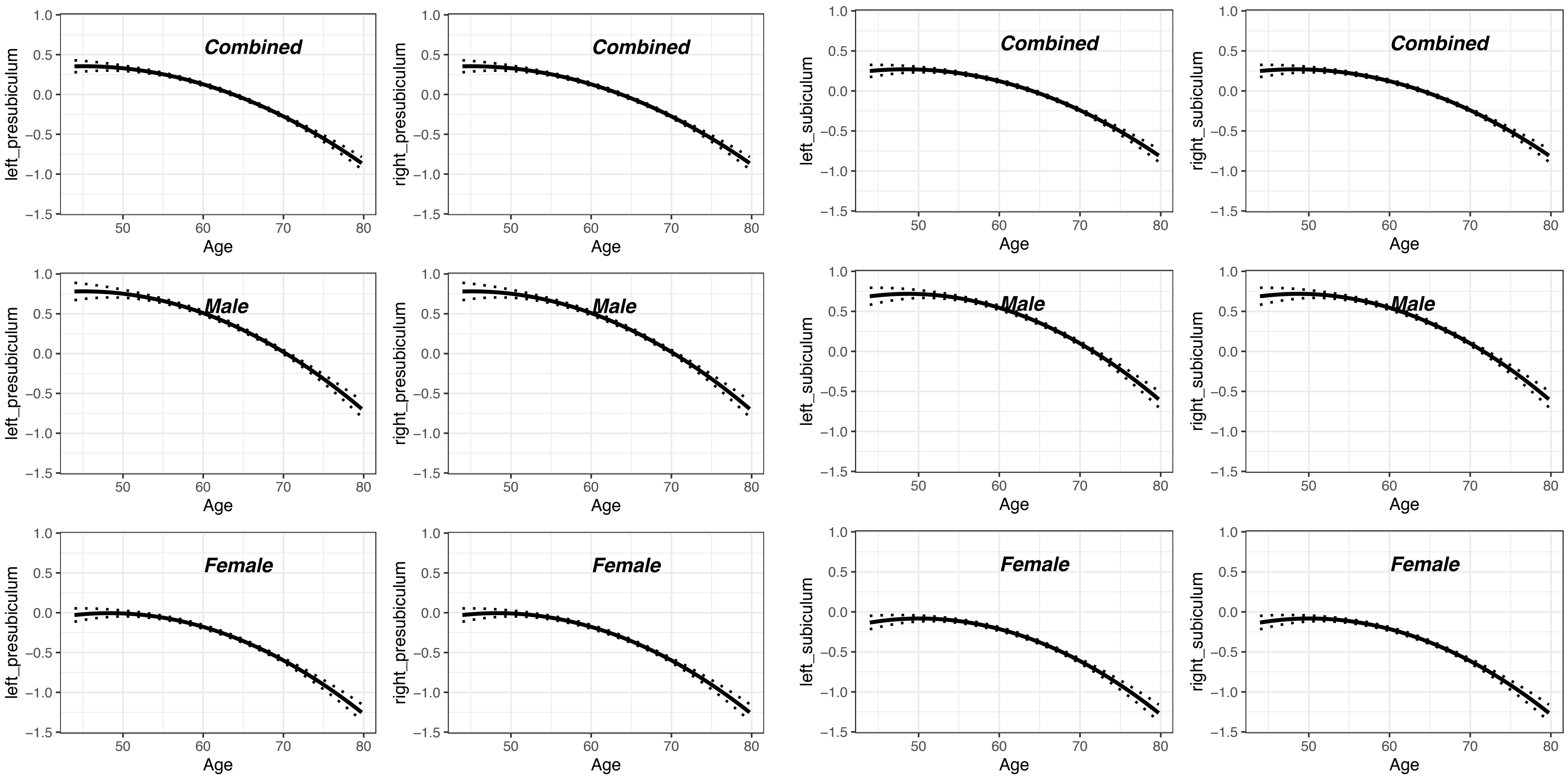

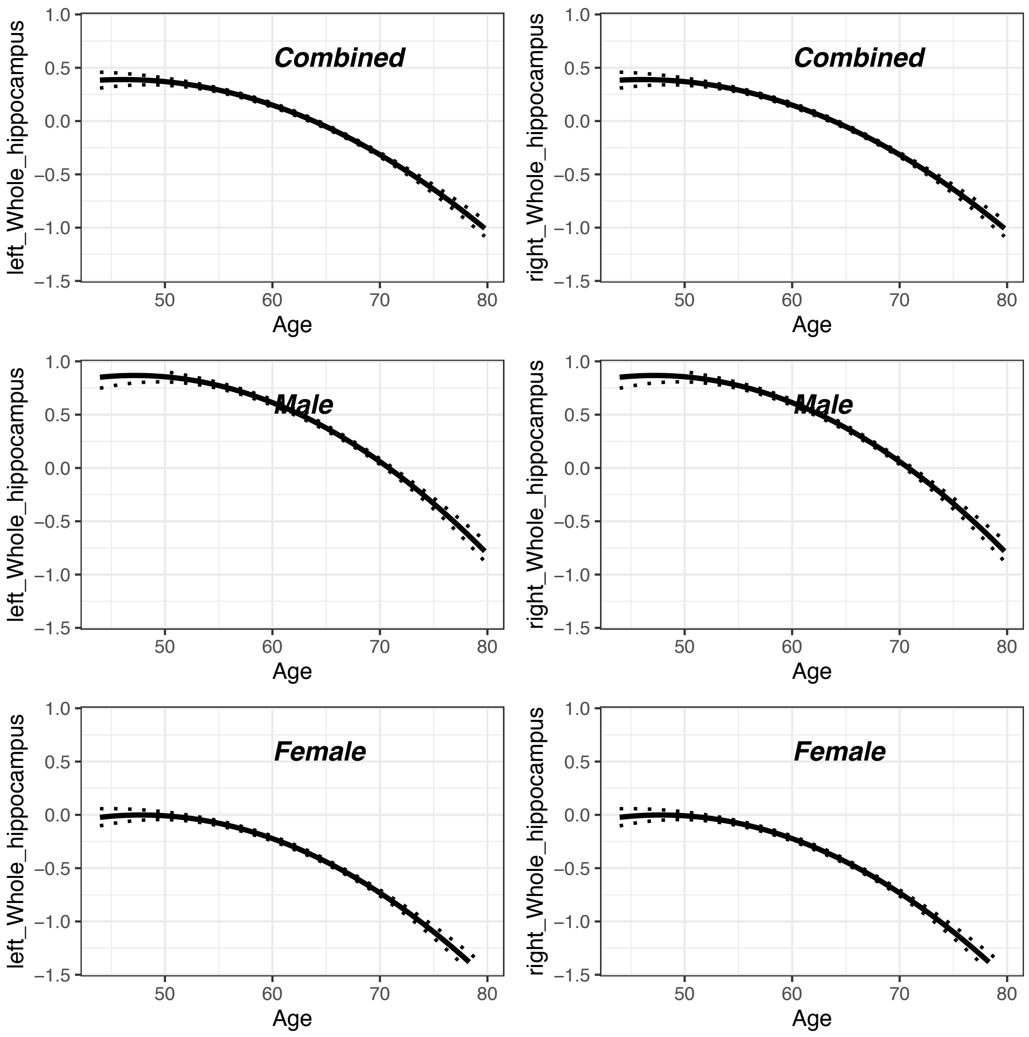

### Supplementary figure 2

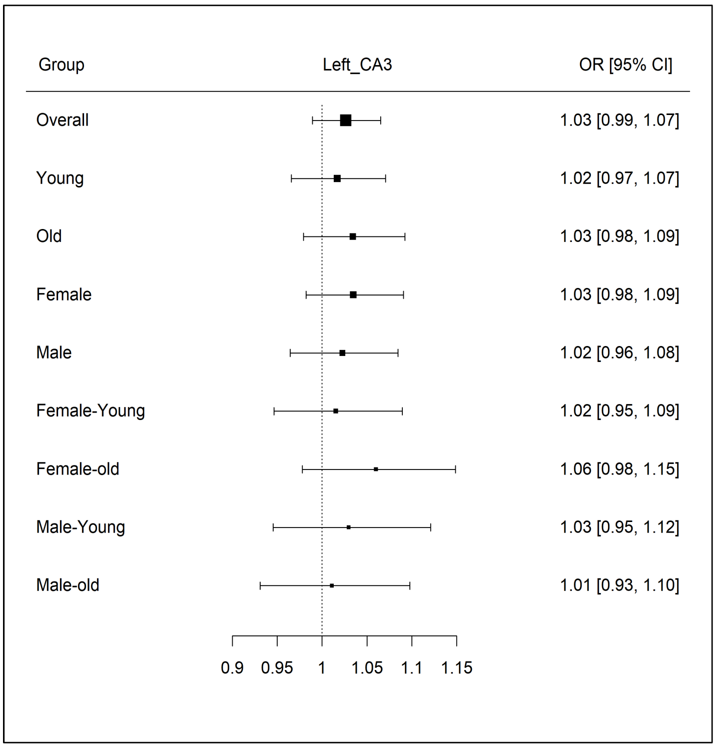

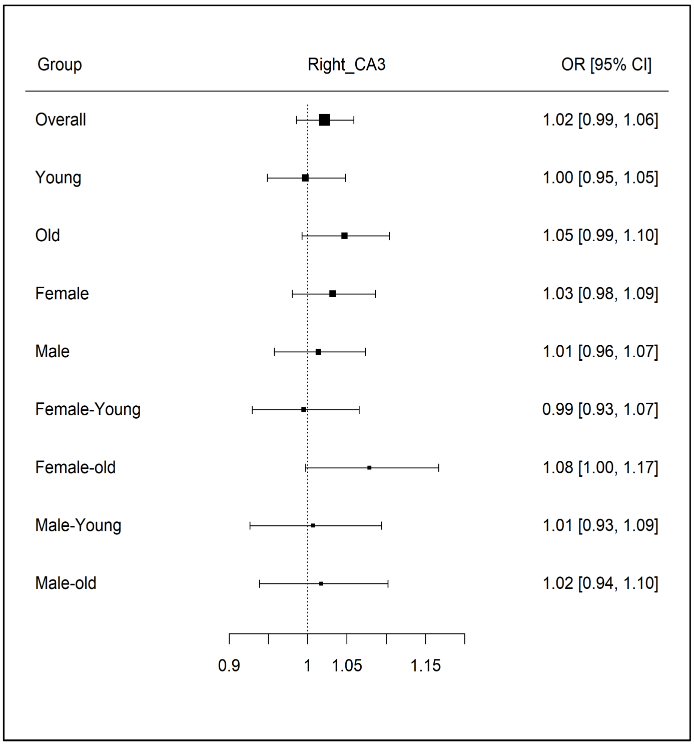

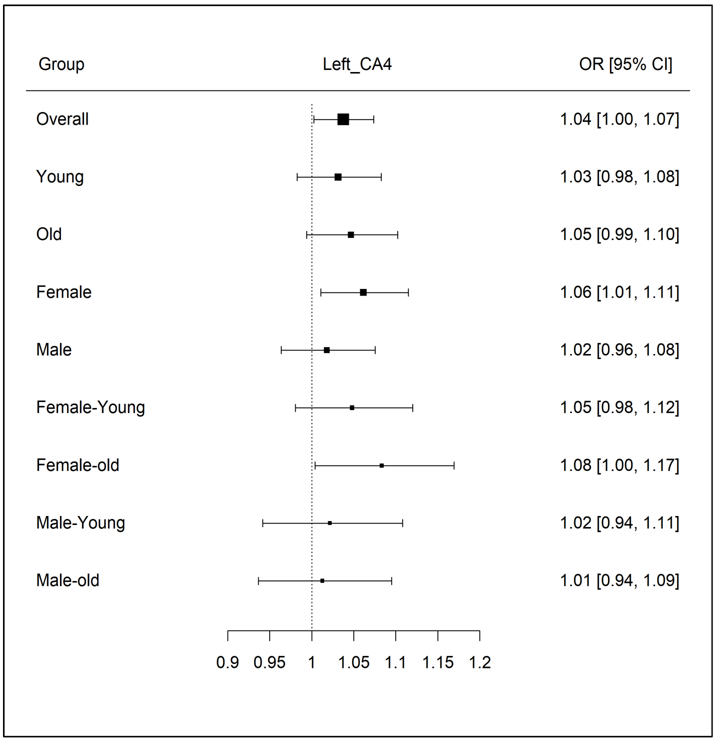

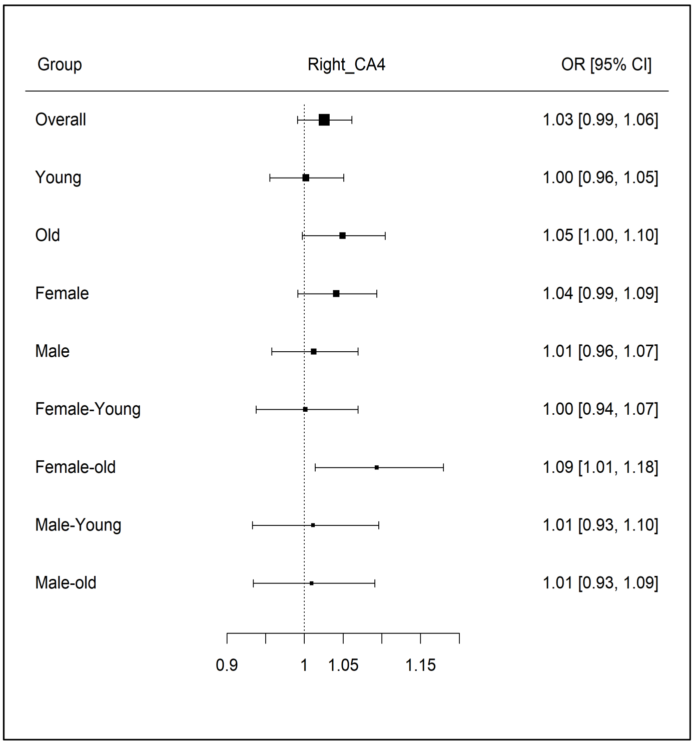

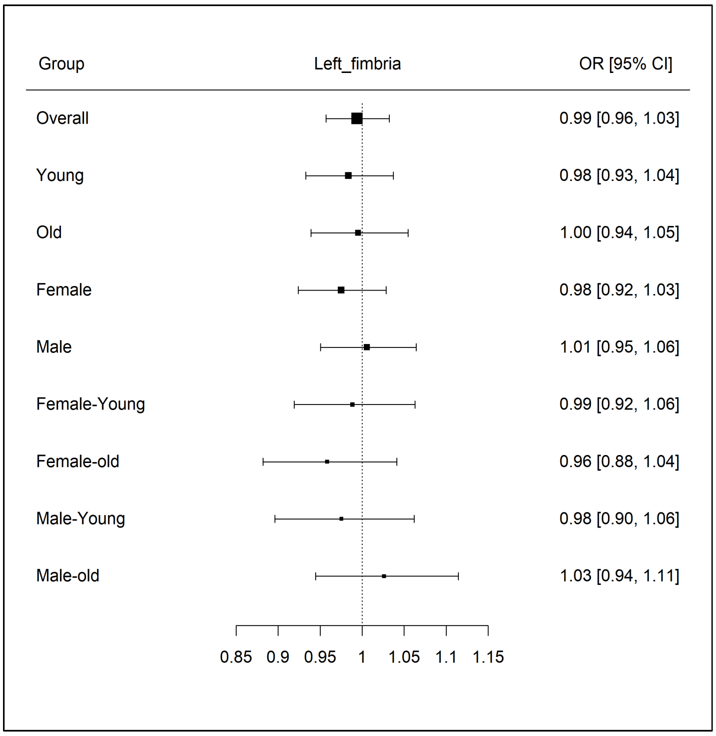

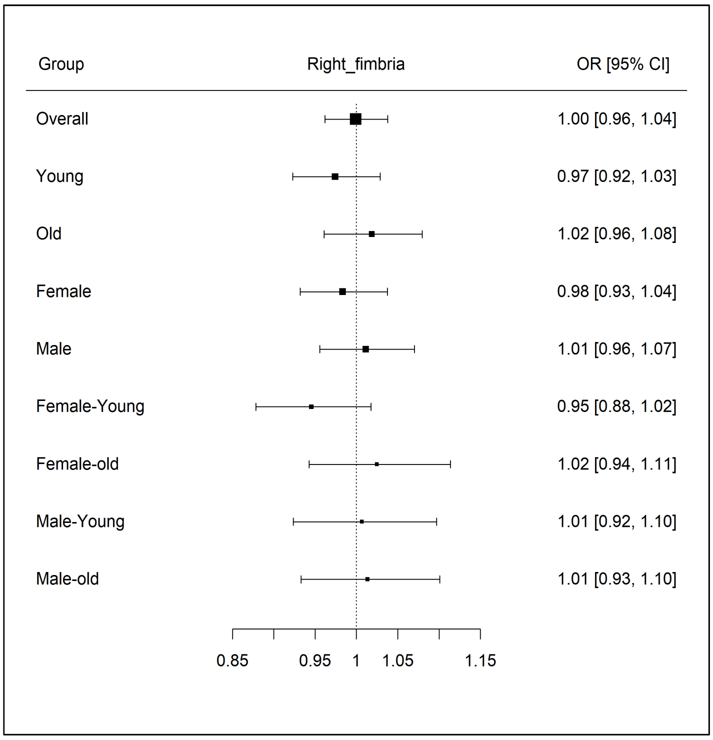

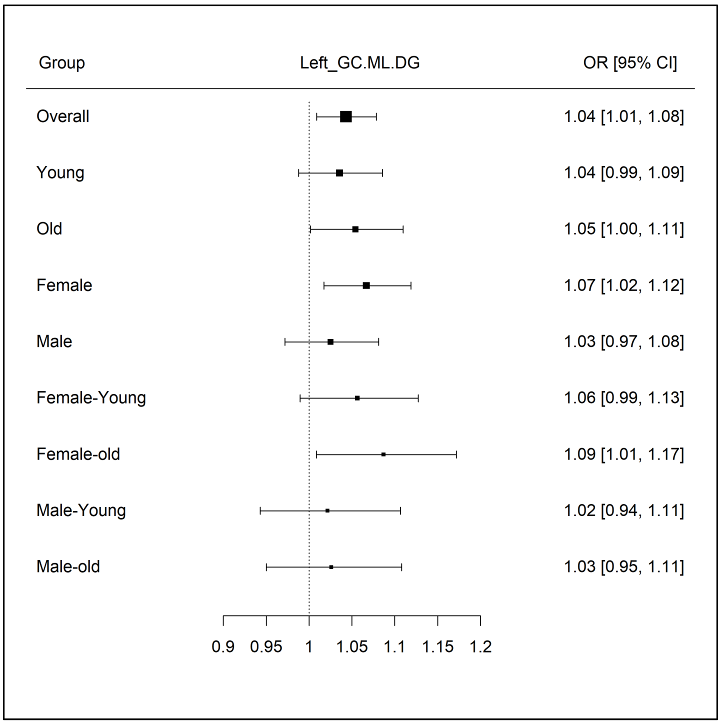

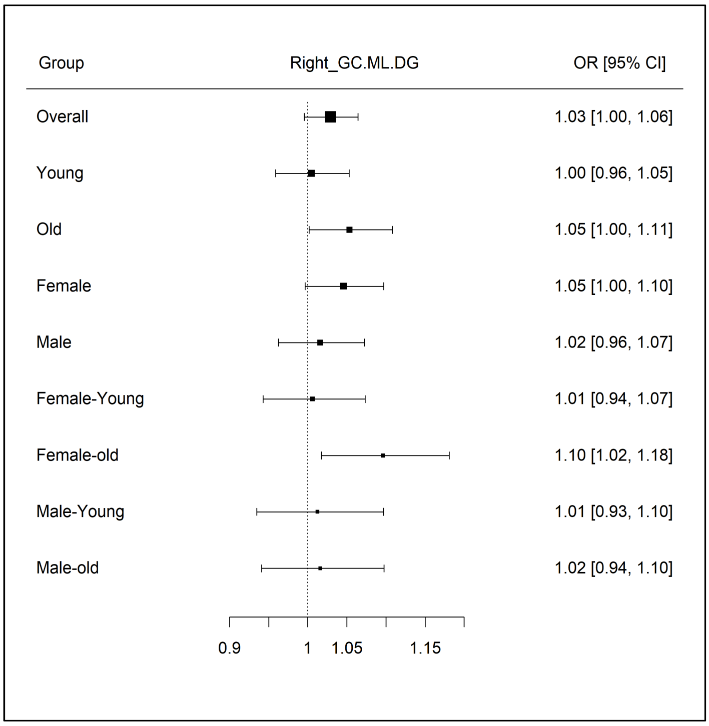

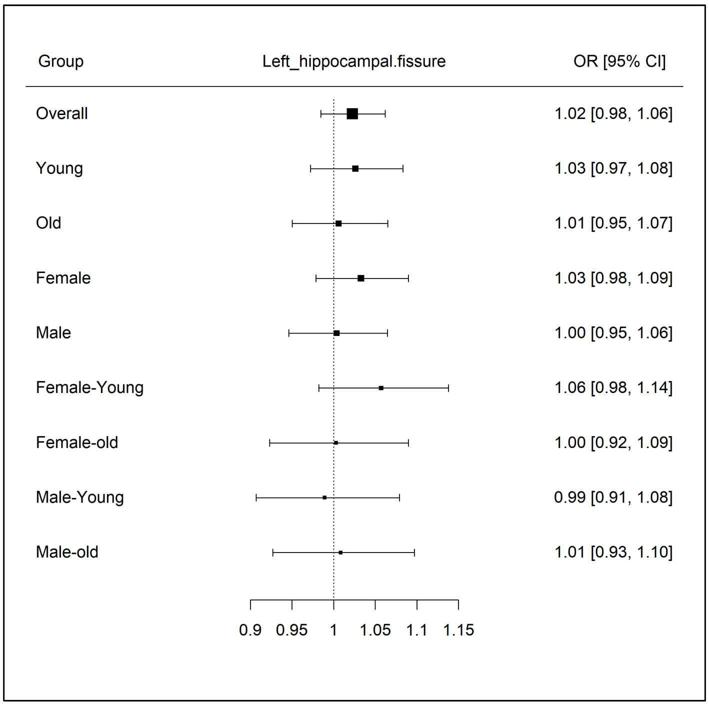

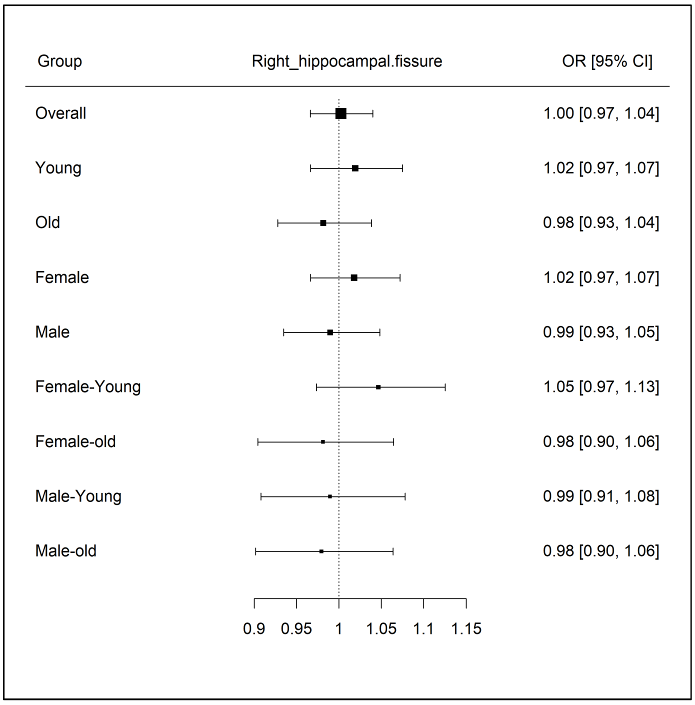

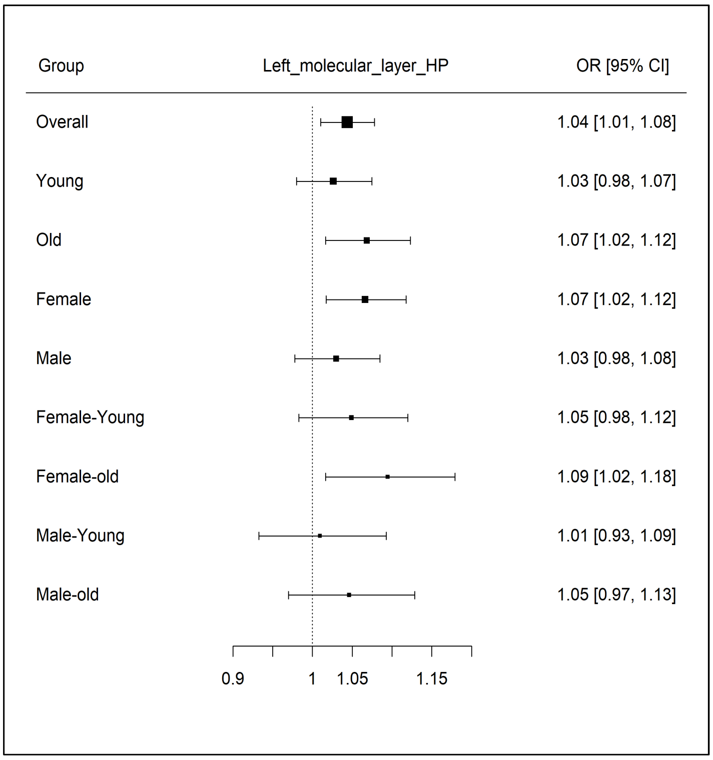

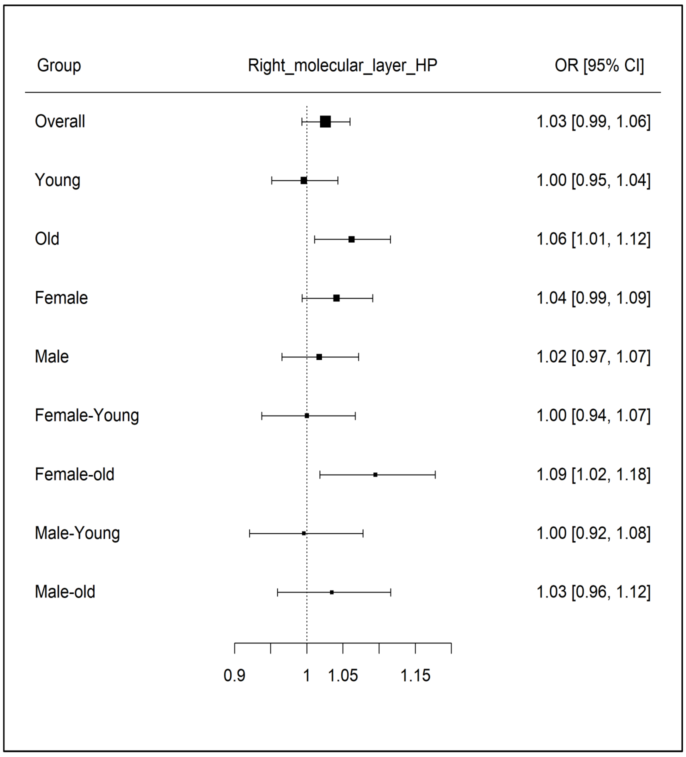

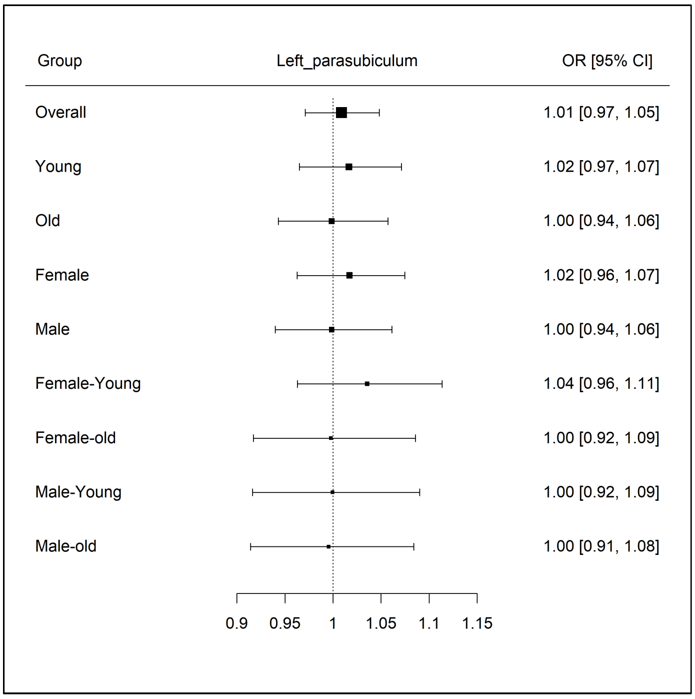

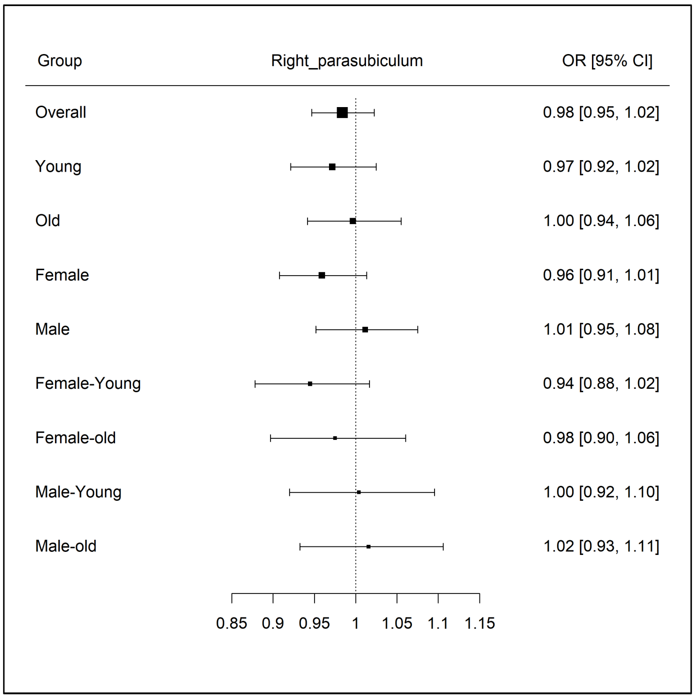

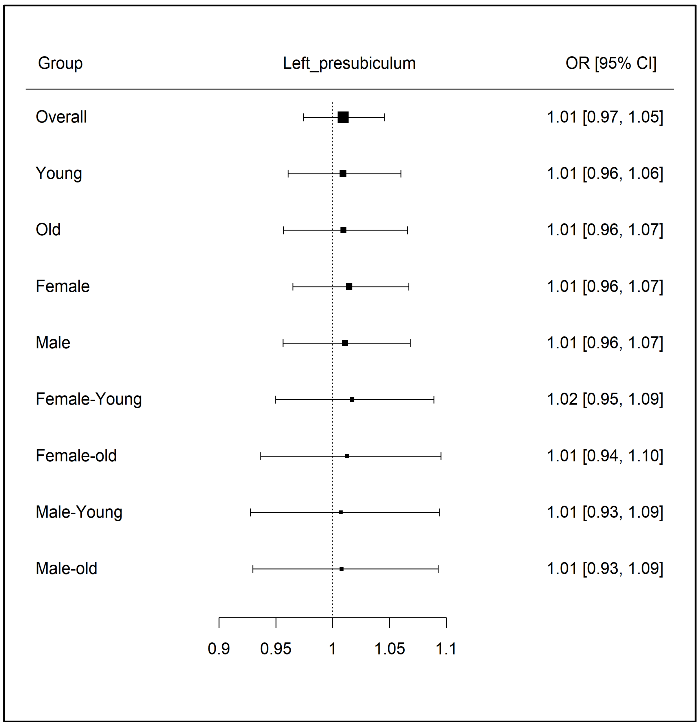

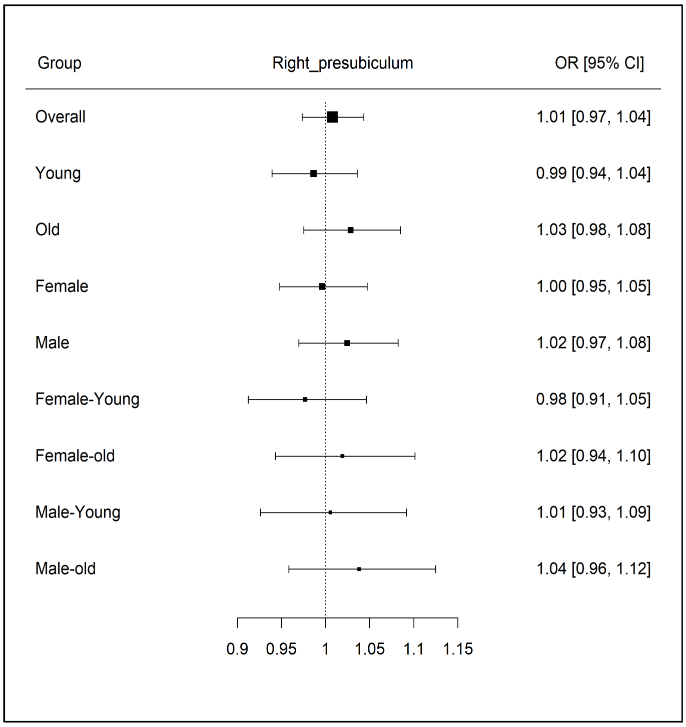

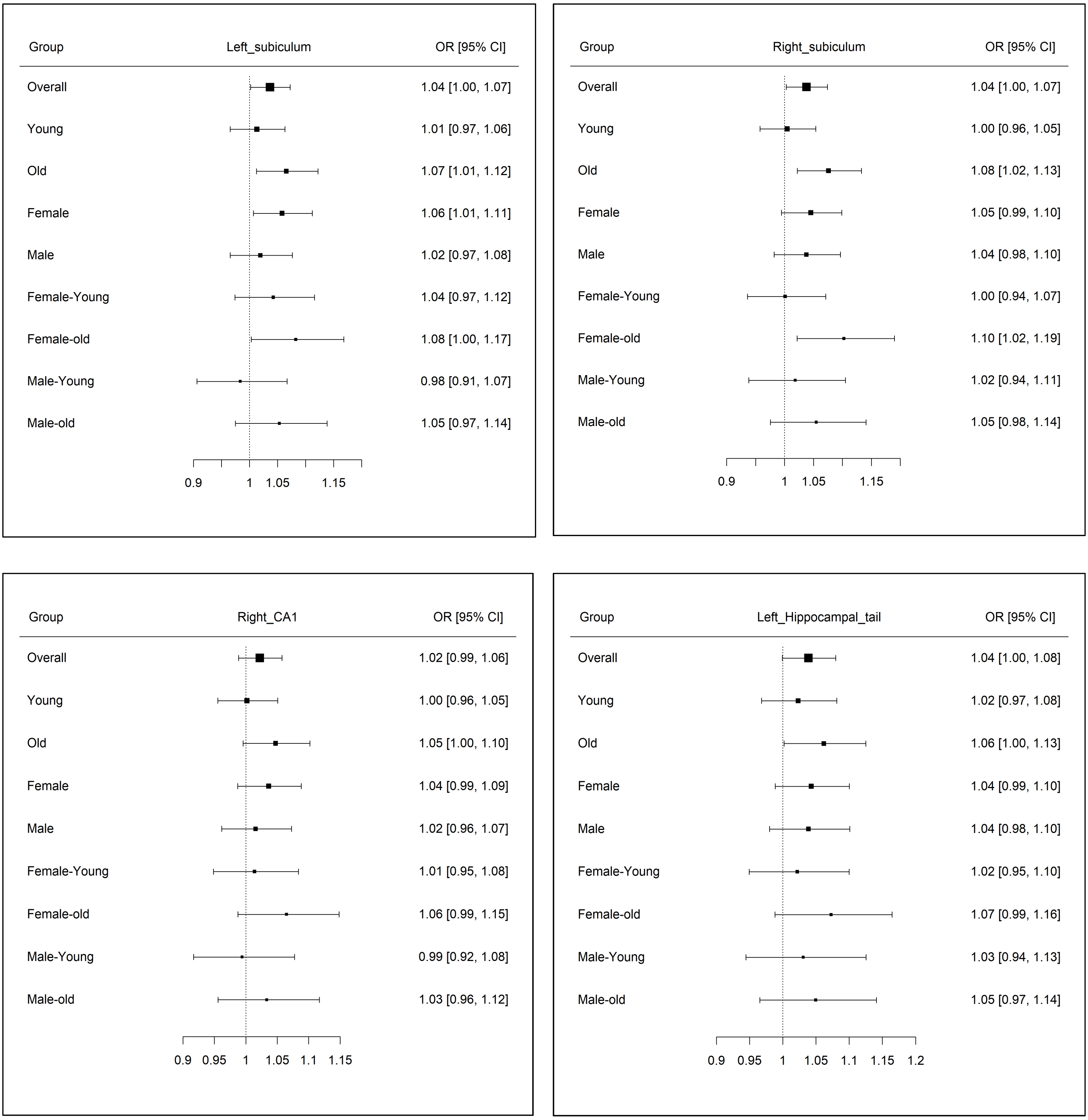

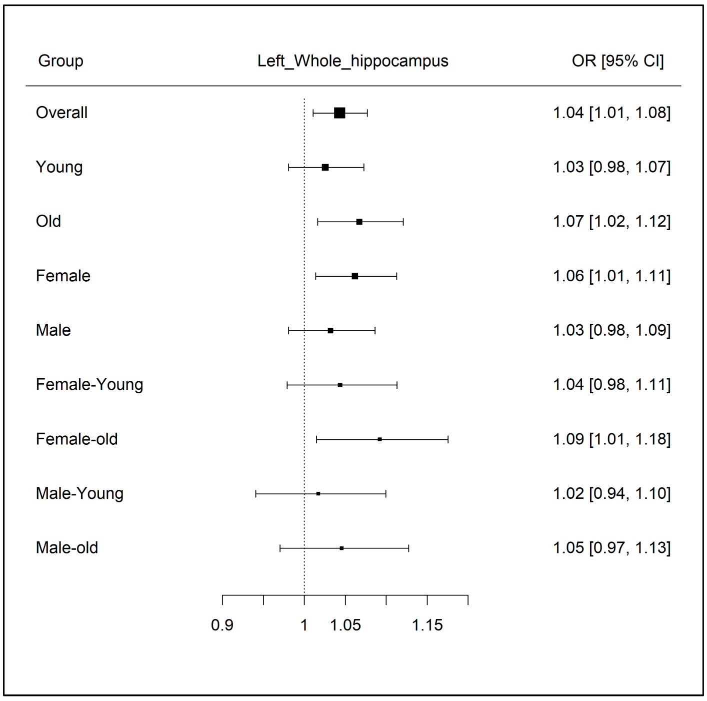

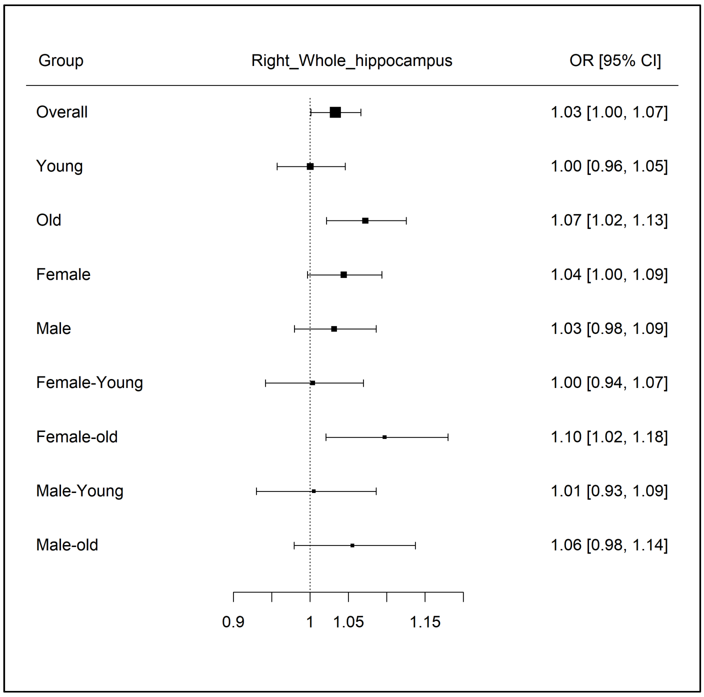
